# Cell-type aware transcriptome-wide association study of mammographic density phenotypes

**DOI:** 10.1101/2025.11.25.25341027

**Authors:** Adriana Sistig, Joseph H. Rothstein, Sinan Zhu, S. Taylor Head, Yung-Han Chang, Ninah Achacoso, Stacey E. Alexeeff, Vignesh Arasu, Tejomay Gadgil, Lawrence D. Gerstley, Laurie R. Margolies, Lori C. Sakoda, Li Shen, Cara L. Smith Gueye, Marvella Villaseñor, Mark Westley, MODE/BCAC Consortia, Arjun Bhattacharya, Robert J. Klein, Laurel A. Habel, Xiaoyu Song, Pei Wang, Weiva Sieh

## Abstract

**Background:** Mammographic density (MD) phenotypes are highly heritable and strongly associated with breast cancer risk. Genetic variants identified by genome-wide association studies (GWAS) explain only a small fraction of the heritability, and the responsible genes remain largely unknown. Transcriptome-wide association studies (TWAS) can improve power and identify genes associated with MD through their genetically regulated gene expression (GReX) levels. However, cell-type heterogeneity in bulk tissue samples can obscure disease associations. Here, we conduct TWAS of MD phenotypes using standard approaches and a new cell-type-aware framework.

**Methods:** The study population included 24,158 women of European ancestry who underwent screening with Hologic (n=20,282) or GE (n=3,876) digital mammography and participated in Kaiser’s Research Program on Genes Environment and Health. Dense area (DA), nondense area (NDA), and percent density (PD) were measured centrally using Cumulus6. Tissue-level gene expression was estimated using standard elastic-net models. Cell-type-specific expression in mammary epithelial, fibroblast, and adipocyte cells were estimated using MiXcan2. Linear regression was used to assess associations of GReX levels with MD phenotypes, adjusted for age at mammography, BMI, and other covariates. Statistical significance was determined by controlling the false-discovery rate at 0.05.

**Results:** A total of 20 genes at 16 independent loci were significantly associated with MD phenotypes, including 6 novel genes at 6 independent loci not found by prior GWAS or TWAS of MD phenotypes. We discovered that one of the novel MD genes, *THBS2-AS1* at 6q27, is also a novel breast cancer susceptibility locus. *THBS2-AS1* expression in mammary tissue was significantly associated with decreased NDA and increased PD, and also with increased breast cancer risk in independent study populations. In comparison, standard TWAS methods identified only 8 MD genes at 7 loci that all were identified by MiXcan2. Additionally, we identified candidate genes for MD phenotypes at 10 known GWAS loci.

**Conclusion:** This TWAS identified novel genes for MD phenotypes and breast cancer risk, and prioritized genes at known GWAS loci that are likely to be causally associated through their expression levels in mammary epithelial, fibroblast, or adipocyte cells. Disentangling the distinct effects of gene expression in different mammary cell types through cell-type-aware analysis can yield new gene discoveries and insights into the biological basis of dense vs. nondense breast tissue.

## INTRODUCTION

Mammary tissue composition is highly variable among women. Visualized on screening mammography, breast composition can range from almost entirely fatty (dark), reflecting a high proportion of adipose tissue, to extremely dense (light), reflecting a high proportion of fibroglandular tissue [1]. Mammographic density (MD) is one of the strongest and most common risk factors for breast cancer. Over 40% of U.S. women have heterogeneously or extremely dense breasts [2]. Women whose breasts appear ≥75% dense have >4-fold increased risks of breast cancer relative to women with little or no dense tissue, independent of other known risk factors [3, 4]. Importantly, whereas the quantitative MD phenotypes, percent density (PD) and dense area (DA) are associated with increased risk, higher nondense area (NDA) is associated with decreased breast cancer risk in both observational [5] and Mendelian randomization studies [6], indicating the disparate roles of different cellular components of mammary tissue in normal breast development and carcinogenesis [7].

Twin studies have shown that DA, NDA, and PD each have heritability estimates >50% [8-10]. Genome-wide association studies (GWAS) have identified variants at 55 independent loci in European ancestry women and 34 loci in African ancestry women associated with one or more MD phenotypes [6, 11-13]. However, altogether these variants explain only a small proportion of the heritability of MD phenotypes [6, 11]. Transcriptome-wide association studies (TWAS) have identified 8 genes, including 2 genes independent from prior GWAS loci, in European ancestry women and 2 genes in African ancestry women that are associated with MD phenotypes through their genetically regulated gene expression (GReX) levels [12, 13]. However, traditional TWAS methods [14, 15] predict GReX at the bulk tissue level and do not account for cell-type heterogeneity, which can obscure disease associations, especially when the most biologically relevant cell type is a minor cell type in the tissue. Cell-type heterogeneity has been shown to be especially important in the mammary tissue context where expression quantitative trait loci (eQTLs) can have different effects in mammary epithelial cells vs. adipocytes [16].

We previously developed MiXcan, a cell-type-aware TWAS framework that predicts cell-type-level GReX and identifies disease-associated genes via combination of cell-type level association signals across different cell types and provides insight into the responsible cell type in the relevant tissue context [17]. Here, we describe MiXcan version 2 (MiXcan2), which employs an ensemble approach to stabilize model selection and improve the robustness of GReX predictions, and perform the first cell-type-aware TWAS of MD phenotypes in 24,158 European ancestry women.

## METHODS

### Study population

The study population included 24,158 European ancestry women from the Kaiser Permanente Northern California (KPNC) Research Program on Genes, Environment and Health (RPGEH [18, 19]) who underwent screening with full-field digital mammography (FFDM) between the ages of 38 and 80 in 2004-2013 at a KPNC clinic. Written informed consent was obtained from all participants. Institutional Review Board approval was obtained from the KPNC, Icahn School of Medicine at Mount Sinai, and University of Texas MD Anderson Cancer Center. Details of the study population and data collection methods have been described previously [6]. Briefly, MD phenotypes were measured by an expert radiological technologist using Cumulus6 on a single craniocaudal view from 20,282 Hologic and 3,876 General Electric (GE) FFDM exams. MD phenotypes (DA, NDA, and PD) were transformed separately within each cohort to attain standard normal distributions and to facilitate meta-analysis and interpretation of effect sizes in standard deviation (SD) units. Genotypes were assayed using the Affymetrix Axiom array with >650,000 variants. In the present study, genotypes were re-imputed using Minimac3 and an expanded reference panel comprising the 1000 Genomes Project Phase 3 and Haplotype Reference Consortium data to improve accuracy for less common variants [20]. Allele dosage effects were estimated using linear regression models adjusted for age at mammography, ln(BMI), the first ten principal components of European ancestry, genotyping reagent kit, and image batch separately in the Hologic and GE cohorts, and the estimates were combined using inverse-variance weighted meta-analysis.

### Training dataset

Models for estimating genetically regulated gene expression (GReX) levels were trained using GTEx v8 genotype and gene expression data for mammary tissue samples from 125 European ancestry women (dbGaP accession number phs000424.v8.p2). An initial set of candidate variants was selected for each gene by extracting all variant-gene pairs with significant expression quantitative trait loci (eQTL) associations for any of 49 tissue types, or significant cell-type interaction eQTL (ieQTL) associations for any of 43 pairs of tissues and cell types [16, 21]. This yielded a total of 37,243 genes and 4,914,536 variants with significant eQTL (n=4,182,730), ieQTL (n=282,079), or both eQTL and ieQTL (n=449,727) associations. We used the European population (n=503) of the 1000 Genomes Project Phase 3 [22] to remove variants with a minor allele frequency (MAF) <1% or pairwise linkage disequilibrium (LD) >0.8 using the PLINK 1.9 --indep-pairwise function with an r^2^ threshold of 0.8 and window size of 20,000, which was larger than the number of variants for any gene, obviating the need for sliding windows [23]. Mammary tissue normalized expression data for 25,849 genes was obtained from GTEx_Analysis_v8_eQTL_expression_matrices.tar (https://gtexportal.org/home/downloads/adult-gtex/qtl). After MAF filtering, LD pruning, and restricting to genes with mammary tissue expression data, the final training set included a total of 25,025 genes with 1-1275 variants per gene (average 91) and 1,142,209 variants with eQTL (n=523,221), ieQTL (n=245,972) or both eQTL and ieQTL (n=373,016) associations.

### Cell-type aware and naive GReX prediction models

MiXcan [17] builds cell-type-level GReX prediction models from bulk tissues, and applies them to genotype data from GWAS to identify disease-associated genes and infer their functional cell types. Specifically, MiXcan first estimates the cell-type proportions for each bulk tissue by integrating prior estimates with cell-type signature gene expression profiles through a joint likelihood function. Then, it builds cell-type-level GReX prediction models by decomposing the observed expression into different cell types and fitting elastic-net models with 10-fold cross validation to jointly estimate the genotype-expression association coefficients for each cell type. Lastly, the cell-type-level GReX prediction models are applied to genotype data from independent GWAS to identify disease-associated genes and signals across cell types are compared to provide insights into the disease-critical cell types.

As a proof-of-concept, the original prediction models developed in MiXcan [17], denoted here as MiXcan1, have several limitations. First, its cell-type proportion estimation framework works only for two cell types, such as epithelial and stromal, while the stromal compartment of mammary tissue consists of biologically distinct components such as fibroblasts and adipocytes that play different roles. Second, MiXcan1 was trained based on the PredictDB (http://predictdb.org) GTEx v8 mammary tissue database, comprising only 6461 genes and 178,698 genetic variants significantly associated with tissue-level expression, potentially excluding a large number of genes and variants associated in a cell-type-specific manner. Third, the elastic-net–based training strategy introduces considerable randomness, especially problematic in small training sets, leading to unstable estimates of genetic effects. To address these limitations, we introduce MiXcan2, an enhanced framework designed to provide multi-cell-type resolution, genome-wide modeling without reliance on PredictDB filters, and substantially improved coefficient stability, with a particular focus on mammographic density, a key intermediate phenotype of breast cancer risk.

We first jointly estimated the cell-type proportions for epithelial cells, fibroblasts, and adipocytes in the 125 GTEx mammary tissue training samples from European ancestry women using BayesDeBulk [24], leveraging the expression levels of cell-type signature genes [25]. With these estimated proportions, MiXcan2 trains cell-type-level prediction models for a total 25,025 genes by focusing on each of the three cell types separately, generating three sets of prediction models in parallel. To overcome the randomness of elastic-net models, MiXcan2 employs a bootstrap strategy which randomly selects 90% of the samples without replacement to fit the model and repeats this process 101 times to create replicates for the ensemble. It is worth noting that each bootstrap sample may result in a cell-type-specific model (CTS) or non-specific (NS) model depending on whether the coefficients differ across cell types. Because coefficients from CTS and NS models are not directly comparable, we employ a consensus voting (>50%) strategy in the ensemble: we first identify the dominant model type (CTS vs NS) across bootstrap replicates, and then average coefficients only within the dominant model type to construct the final prediction model. MiXcan2 ensemble models were estimated using the MiXcan2_Ensemble.R function.

For comparison, we used the PrediXcan [14] approach to train standard tissue-level GReX prediction models (hereafter referred to as “PrediXcan” models) using the same training set of 125 GTEx mammary tissue samples that was used to train the MiXcan2 models. PrediXcan models were estimated using the MiXcan2_model.R function specifying NS models only. All models were adjusted for covariates that were used in GTEx eQTL analyses, including age, sequencing platform, library construction protocol, 5 genomic principal components, and 15 Probabilistic Estimation of Expression Residuals (PEER [26]) factors to account for unmeasured confounders. Genes with GReX prediction models that had a coefficient of determination (cross-validated adjusted R^2^) value greater than 0 were retained in downstream analyses.

### Evaluation of GReX prediction models

We evaluated the performance accuracy of MiXcan2 and PrediXcan GReX models trained using GTEx v8 mammary tissue data in an independent dataset of 102 tumor-adjacent normal mammary tissue samples from European ancestry women in The Cancer Genome Atlas (TCGA). TCGA gene expression data were re-processed using a pipeline parallel to GTEx, as previously described [17]. Genotype data was obtained from dbGaP (study accession phs000178) and imputed with the TOPMed imputation server. Imputed genetic variants were required to have an imputation accuracy metric (R^2^) of at least 0.3. Cell-type proportions (𝛑) for TCGA bulk mammary tissue samples were jointly estimated for epithelial cells, fibroblasts, and adipocytes using BayesDeBulk [24] and the expression levels of cell-type signature genes [25]. Genes with GReX prediction models that had a coefficient of determination (cross-validated adjusted R^2^) value greater than 0, and at least one SNP predictor in the TCGA genotype data were assessed. Mammary tissue-level expression of cell-type-specific genes was computed using a weighted mean, Ŷ = (𝛑*Ŷ_1_) + ((1-𝛑)*Ŷ_2_), of the cell-type-level predictions. Similarity between predicted and observed expression levels were assessed with Pearson correlation coefficients.

### Transcriptome-wide association study (TWAS) analyses

To assess the association of gene expression in the mammary tissue context with MD phenotypes, we conducted cell-type-aware TWAS using MiXcan2 GReX models for mammary epithelial, fibroblast, and adipocyte cells and standard TWAS using mammary tissue models. Genetic variants that were not genotyped in the RPGEH study population were excluded when computing predicted gene expression levels. Linear regression models were fit to estimate the cell-type specific or non-specific genetic effect, adjusting for age at mammography, BMI, genotyping reagent kit, and 10 principal components of ancestry. The effect size and standard error were obtained separately for women with GE or Hologic mammograms, and inverse-variance weighted meta-analysis was used to obtain the combined meta-analytic effect size, standard error, and p-value. For cell-type-specific models, the aggregated Cauchy association test (ACAT [27]) was applied to combine the cell-specific p-values into a composite tissue-level p-value that was used to test for statistical significance. The Benjamini-Hochberg adjustment procedure was applied to control the false discovery rate and significant associations were determined by an adjusted p-value < 0.05. Quantile-quantile plots and λ_1000_ [28] were used to assess genomic inflation. λ_1000_ represents the value of the standard genomic control factor (λ [29]) that would be expected from an otherwise equivalent study with a sample size of 1000.

### Associations with breast cancer risk

To determine whether significant genes in our TWAS were previously associated with breast cancer risk, we examined overlap with previously reported breast cancer GWAS loci and TWAS genes. We downloaded genome-wide summary statistics for two Breast Cancer Association Consortium (BCAC) GWAS [30, 31] from <https://www.ccge.medschl.cam.ac.uk/breast-cancer-association-consortium-bcac/data-data-access/summary-results>, and used results from the largest available meta-analysis of overall breast cancer risk from each study. We identified genome-wide significant variants (p < 5*10^-8^) within 1 Mb of each MD gene, and recorded the most statistically significant variant. We also examined whether significant MD genes were previously associated with breast cancer risk through their GReX levels by looking up the results in recent large breast cancer TWAS [17, 32-39].

We tested the associations of significant MD genes with overall breast cancer risk in an independent population of 133,384 case and 113,789 control women of European ancestry [31] using GWAS summary statistics and the same GReX models as in the discovery population. S-MiXcan [40] was used to evaluate cell-type-specific and non-specific epithelial, fibroblast, and adipocyte models, and the S-PrediXcan [41] approach implemented using S-MiXcan software was used to evaluate mammary tissue-level GReX prediction models. Genetic variants that were present in GReX prediction models, GWAS datasets, and the 1000 Genomes Project Phase 3 European (n=633) population [22] were harmonized by flipping effect directions when reference and alternate alleles were reversed between datasets and removing indels and ambiguous (palindromic) SNPs. We used PLINK 2.0 [23] to compute linkage disequilibrium matrices in the 1000 Genomes population and applied S-MiXcan to compute cell-type level Z-scores and p-values, and the ACAT-combined tissue-level composite p-values. Statistical significance was determined by a tissue-level p or ACAT-combined p < 0.0025 applying a Bonferroni correction for the 20 MD genes tested.

### Replication analyses

To determine whether significant genes in our TWAS were previously associated with MD phenotypes, we examined overlap with previously reported MD GWAS loci and TWAS genes. We downloaded all genome-wide significant (p<5*10^-8^) associations with the “mammographic density measurement” trait (EFO_0005941) from the GWAS Catalog [42] on August 18, 2025 and recorded the earliest published variant within 1 Mb of each gene. We also examined whether significant MD genes were previously associated with MD phenotypes through their GReX levels by looking up the results when available in prior TWAS in European ancestry [12] and African ancestry [13] women. We recorded genes that met the authors’ transcriptome-wide significance threshold, and the significance levels and directions of effect when reported for other genes.

We also performed replication analyses in an independent population of up to 27,900 European ancestry [12] using GWAS summary statistics for DA, NDA, and PD downloaded from <https://www.ccge.medschl.cam.ac.uk/breast-cancer-association-consortium-bcac/data-data-access/summary-results/gwas-summary-results-1>. S-MiXcan [40] was used to evaluate cell-type-specific and non-specific epithelial, fibroblast, and adipocyte GReX models, and the S-PrediXcan [41] approach implemented using S-MiXcan software was used to evaluate mammary tissue-level GReX prediction models as described above.

### Colocalization

Evidence of colocalization, defined as the presence of at least one genetic variant that is causally associated with both gene expression and the phenotype, was assessed using eCAVIAR version 2.2 [43]. We computed genome-wide association statistics for DA, NDA, and PD in the study population of 24,158 European ancestry women using previously described methods [6]. We downloaded GTEx v8 eQTL summary statistics for the association of all variants within 1 Mb of the transcription start site with gene expression levels in bulk mammary tissue samples from 329 men and women of European ancestry <https://console.cloud.google.com/storage/browser/gtex-resources/GTEx_Analysis_v8_QTLs/GTEx_Analysis_v8_EUR_eQTL_all_associations>. Pairwise linkage disequilibrium (*r*) in the 1000 Genomes Project Phase 3 European sample (n=503) was computed using PLINK 2.0 [23] for all variants that had MD GWAS, GTEx eQTL, and 1000 Genomes Project data. Colocalization analyses were performed separately for each significant gene and its associated MD phenotype(s) to estimate the colocalization posterior probability (CLPP) for each genetic variant, specifying a maximum of two causal variants. A CLPP threshold of 0.01 was considered evidence of colocalization [43].

### Data availability

The data used in this study are available from the following sources: GTEx v8 (dbGaP accession number phs000424.v8.p2 <https://www.ncbi.nlm.nih.gov/projects/gap/cgi-bin/study.cgi?study_id=phs000424.v8.p2>); TCGA (dbGaP accession number phs000178.v8.p7 <https://www.ncbi.nlm.nih.gov/projects/gap/cgi-bin/study.cgi?study_id=phs000178.v8.p7>); and the Breast Cancer Association Consortium (BCAC) (https://www.ccge.medschl.cam.ac.uk/breast-cancer-association-consortium-bcac/data-data-access/summary-results).

Kaiser Permanente Research Program on Genes, Environment, and Health (RPGEH) and the Genetic Epidemiology Research on Adult Health and Aging (GERA) genotype data are available from dbGaP accession phs000674.v3.p3 (https://www.ncbi.nlm.nih.gov/projects/gap/cgi-bin/study.cgi?study_id=phs000674.v3.p3). This includes individuals who consented to having their data shared with dbGaP. Access to the data used in this study may be obtained by application to the Kaiser Permanente Research Bank (kp.org/researchbank/researchers).

### Code availability

The MiXcan2 R package is publicly available at: https://github.com/songxiaoyu/MiXcan2/. The S-MiXcan R package is publicly available at: https://github.com/songxiaoyu/SMiXcan.

## RESULTS

### Cell-type aware and naive TWAS

Cell-type-aware TWAS was conducted using MiXcan2 epithelial, fibroblast, and adipocyte cell GReX prediction models available for 14,248 (5608 CTS and 8640 NS), 13,604 (5745 CTS and 7859 NS), and 12,206 (4602 CTS and 7604 NS) genes, respectively. Standard TWAS was conducted using PrediXcan mammary tissue GReX prediction models available for 7629 genes. In the training set of 125 European ancestry women with mammary tissue data in GTEx v8, the mean cross-validated coefficient of determination of MiXcan2 epithelial, fibroblast, and adipocyte cell models, respectively, was generally higher for CTS (0.28, 0.23, 0.18) than NS (0.18, 0.17, 0.17) genes, and also higher than PrediXcan tissue-level GReX models (0.08). In the evaluation set of 102 European ancestry women with tumor-adjacent normal mammary tissue samples in TCGA, the correlation of the predicted and observed tissue level gene expression also was generally higher for CTS than for NS models, and was similar for MiXcan and PrediXcan for NS genes (**Supplementary Figure 1**). The genomic inflation factor λ_1000_ ranged from 1.004 to 1.011 in cell-type-aware TWAS and 1.010 to 1.022 in standard TWAS of DA, NDA and PD indicating little evidence of uncontrolled population substructure in the RPGEH study population of 24,158 European ancestry women (**Supplementary Figure 2**).

In total, 20 genes at 16 independent loci were significantly associated with DA, NDA, or PD at the Benjamini-Hochberg false discovery rate (FDR) of 0.05 (**Figure 1**, **Table 1**, **Supplementary Table 1**). Non-specific GReX models estimated by MiXcan2 detected 10 genes at 10 loci associated with DA, NDA, or PD, among which 6 genes also were detected by standard TWAS and 4 genes (*AQP7, ILK, CCND1, ZNF500*) were detected by MiXcan2 only. Both approaches detected a non-specific association of *THBS2-AS1* with NDA, but PrediXcan also detected an association of *THBS2-AS1* with PD that was missed by MiXcan2. Cell-type-specific GReX models estimated by MiXcan2 detected 10 genes at 8 loci associated with DA, NDA, or PD, among which only 2 genes (*VPS13D* and *NTN5*) also were detected using standard TWAS models that assume homogeneous effects across all cell types in the tissue. These results show that allowing for cell-type heterogeneity can improve the discovery of disease-associated genes.

**Figure 1.**
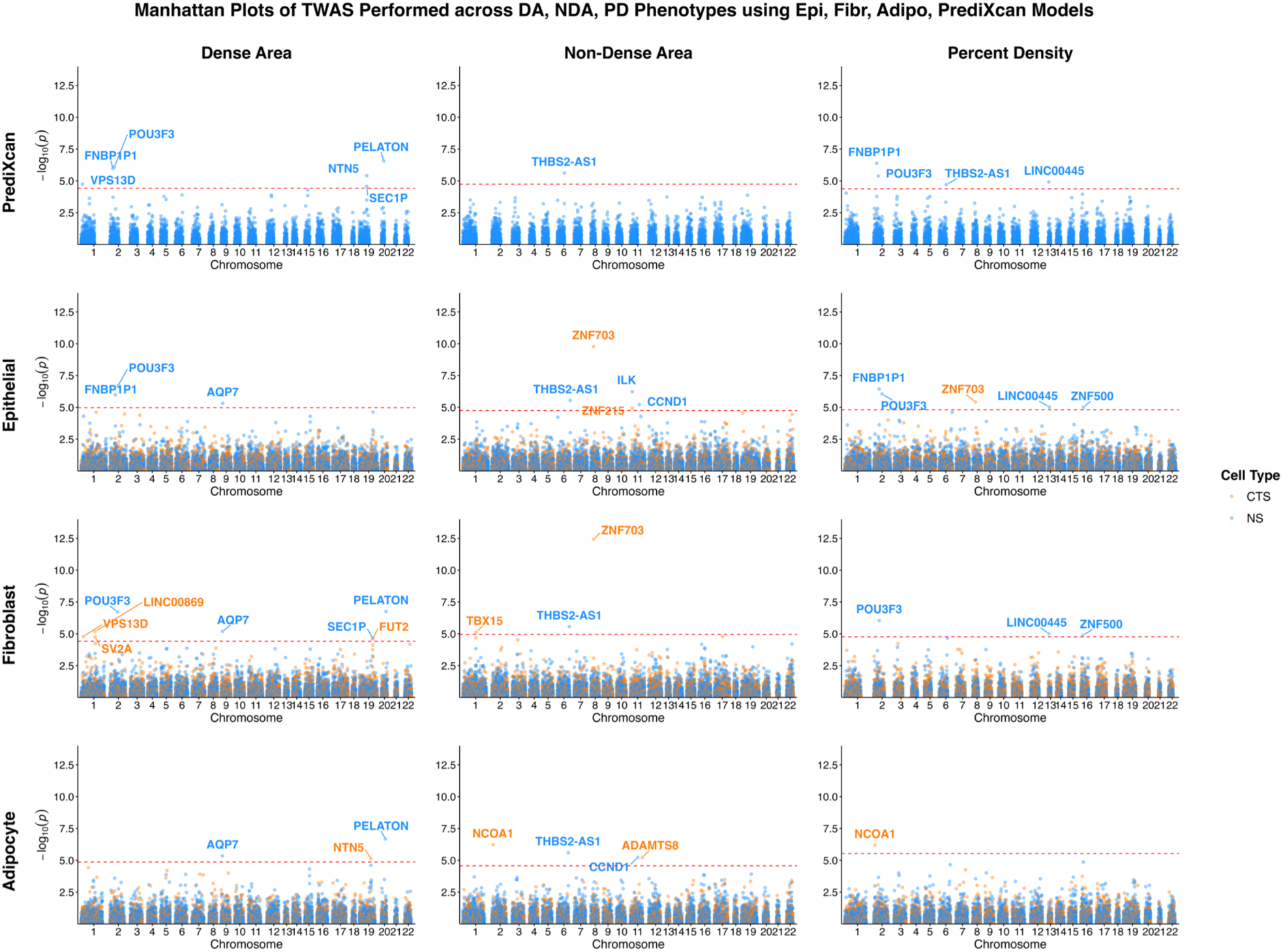
Manhattan plot of cell-type specific (orange) and nonspecific (blue) genetic associations with dense area (DA), nondense area (NDA) and percent density (PD) in standard TWAS and cell-type-aware TWAS using epithelial, fibroblast, or adipocyte cell models.

**Table 1.** Genes significantly associated with mammographic density phenotypes through their genetically regulated gene expression at the mammary tissue or cell-type-specific levels.

| Cytoband | Gene | Start Position<sup>a</sup> | PrediXcan Mammary Tissue P | MiXcan Non-Specific Model P<sup>b</sup> | MiXcan Cell-Type-Specific Associations |  |  |  |
| --- | --- | --- | --- | --- | --- | --- | --- | --- |
|  |  |  |  |  | Model | Overall P | Cell of Interest | Other Cells |
| Dense Area |  |  |  |  |  |  |  |  |
| 1p36.22 | VPS13D | 1:12,230,030 | 1.8e-05 (+) | — | Fibroblast | 1.6e-05 | 8.1e-06 (+) | 2.7e-03 (+) |
| 1q21.2 | LINC00869 | 1:149,655,731 | — | — | Fibroblast | 6.6e-06 | 6.6e-06 (-) | 6.6e-06 (+) |
|  | SV2A | 1:149,903,318 | — | — | Fibroblast | 1.8e-05 | 4.7e-01 (-) | 9.2e-06 (-) |
| 2p13.1 | FNBP1P1 | 2:74,120,680 | 1.1e-06 (-) | 1.0e-06 (-) | — | — | — | — |
| 2q12.1 | POU3F3 | 2:104,853,287 | 7.9e-07 (+) | 1.8e-07 (+) | — | — | — | — |
| 9p13.3 | AQP7 | 9:33,383,179 | — | 4.4e-06 (-) | — | — | — | — |
| 19q13.33 | NTN5 | 19:48,661,407 | 4.0e-06 (-) | — | Adipose | 7.5e-06 | 2.7e-01 (-) | 3.7e-06 (-) |
|  | SEC1P | 19:48,673,174 | 2.7e-05 (-) | 2.1e-05 (-) | — | — | — | — |
|  | FUT2 | 19:48,695,957 | — | — | Fibroblast | 2.4e-05 | 3.1e-05 (+) | 2.0e-05 (+) |
| 20q13.13 | PELATON | 20:50,233,416 | 2.7e-07 (-) | 1.8e-07 (-) | — | — | — | — |
| Non-Dense Area |  |  |  |  |  |  |  |  |
| 1p12 | TBX15 | 1:118,883,046 | — | — | Fibroblast | 7.3e-06 | 3.7e-06 (+) | 7.2e-04 (+) |
| 2p23.3 | NCOA1 | 2:24,490,992 | — | — | Adipose | 6.0e-07 | 9.7e-02 (+) | 3.0e-07 (+) |
| 6q27 | THBS2-AS1 | 6:169,213,118 | 2.4e-06 (-) | 2.9e-06 (-) | — | — | — | — |
| 8p11.23 | ZNF703 | 8:37,695,782 | — | — | Epithelial | 1.7e-10 | 8.9e-11 (+) | 1.2e-09 (+) |
|  |  | 8:37,695,782 | — | — | Fibroblast | 3.6e-13 | 1.8e-13 (+) | 3.5e-05 (+) |
| 11p15.4 | ILK | 11:6,603,708 | — | 6.0e-07 (+) | — | — | — | — |
|  | ZNF215 | 11:6,926,373 | — | — | Epithelial | 1.1e-05 | 9.1e-06 (+) | 1.5e-05 (-) |
| 11q13.3 | CCND1 | 11:69,641,143 | — | 5.9e-06 (-) | — | — | — | — |
| 11q24.3 | ADAMTS8 | 11:130,404,923 | — | — | Adipose | 6.0e-06 | 3.0e-06 (-) | 2.7e-04 (-) |
| Percent Density |  |  |  |  |  |  |  |  |
| 2p23.3 | NCOA1 | 2:24,490,992 | — | — | Adipose | 6.5e-07 | 7.7e-01 (-) | 3.2e-07 (-) |
| 2p13.1 | FNBP1P1 | 2:74,120,680 | 4.1e-07 (-) | 3.6e-07 (-) | — | — | — | — |
| 2q12.1 | POU3F3 | 2:104,853,287 | 4.2e-06 (+) | 8.9e-07 (+) | — | — | — | — |
| 6q27 | THBS2-AS1 | 6:169,213,118 | 2.0e-05 (+) | — | — | — | — | — |
| 8p11.23 | ZNF703 | 8:37,695,782 | — | — | Epithelial | 3.7e-06 | 2.2e-06 (-) | 1.2e-05 (-) |
| 13q13.3 | LINC00445 | 13:35,694,965 | 1.2e-05 (-) | 9.6e-06 (-) | — | — | — | — |
| 16p13.3 | ZNF500 | 16:4,748,239 | — | 1e-07 (-) | — | — | — | — |

a. Gene start positions from GRCh38 (Genome Reference Consortium Human Build 38), Ensembl Genes 114, GENCODE 48 using Ensembl BioMart (August 2025).
b. The most significant p-value among non-specific MiXcan models, which differ slightly due to randomness in the ensemble model estimation procedure.

Among the 7 loci with 10 genes that were significantly associated with DA, 4 loci were reported in prior GWAS [6, 44] including 2 genes, *MTMR11* at 1q21.2 and *PELATON/SMIM25* at 20q13.13, that also were reported by a prior TWAS [12] (**Table 1**, **Supplementary Table 2**). The 3 novel DA loci were *VPS13D* at 1p36.22, *POU3F3* at 2q12.1, and *AQP7* at 9p13.3. Among the 7 loci with 8 genes that were significantly associated with NDA, 6 loci were reported in prior GWAS [6, 11]. We discovered a novel locus, *THBS2-AS1* at 6q27, that was significantly associated with both NDA and PD. Among the 7 loci with 7 genes that were significantly associated with PD, 2 loci (*FNBP1P1* at 2p13.1 and *POU3F3* at 2q12.1) also were associated with DA and 3 loci (*NCOA1* at 2p23.3, *THBS2-AS1* at 6q27, and *ZNF703* at 8p11.23) also were associated with NDA. Four PD loci were reported in prior GWAS [6, 11]. The 3 novel PD loci were *POU3F3* at 2q12.1, *THBS2-AS1* at 6q27, and *ZNF500* at 16p13.3.

### Associations with breast cancer

We reviewed the literature to determine whether significant MD loci identified in this TWAS were previously associated with overall breast cancer risk (**Supplementary Table 2**). Eight of the 16 MD loci, including 2 DA (1q21.2 and 20q13.13), 5 NDA (1p12, 2p23.3, 8p11.23, 11q13.3, and 11q24.3), and 3 PD (2p23.3, 8p11.23, and 16p13.3) loci were reported in prior GWAS of breast cancer [30, 31], including 3 genes (*NCOA1* at 2p23.3, *ZNF703* at 8p11.23, and *CCND1* at 11q13.3) that were associated with overall breast cancer risk in prior TWAS of breast cancer [17, 38].

We also tested the associations of the 20 MD genes with overall breast cancer risk in 133,384 breast cancer cases and 113,789 control women of European ancestry [31] using the same GReX prediction models as in the discovery TWAS (**Table 2**, **Supplementary Table 3**). Four MD genes at 4 distinct loci also were significantly associated with breast cancer risk at the Bonferroni-corrected threshold of 0.0025. *NCOA1* expression, which was positively associated with NDA and inversely associated with PD in mammary adipocyte models, also was inversely associated with breast cancer risk (p = 2.1e-04) in concordance with the directions of association with MD phenotypes. *THBS2-AS1* expression, which was inversely associated with NDA and positively associated with PD in non-specific mammary tissue models, also was positively associated with breast cancer risk (p = 8.7e-07) as expected. *ZNF703* expression, which was positively associated with NDA in mammary fibroblast and epithelial cell models and inversely associated with PD in epithelial models, was positively associated with breast cancer risk in fibroblast and epithelial cell models. There was evidence of cell-type heterogeneity of the association of *ZNF703* expression with breast cancer risk with stronger associations found in mammary fibroblasts (p = 1.4e-21) than other cell types, reflecting the complex genetic regulation at this locus. *SEC1P* expression, which was inversely associated with DA in non-specific mammary-tissue models, was newly positively associated with breast cancer risk (p = 2.7e-05). Future studies are needed to confirm whether *SEC1P* is a breast cancer susceptibility locus.

**Table 2.** Mammographic density genes associated with breast cancer^a^ through their genetically regulated gene expression at the mammary tissue or cell-type-specific levels.

| Cytoband | Gene | Start Position | Associated Density Phenotype(s) | S-PrediXcan Mammary Tissue P | S-MiXcan Non-Specific Model P<sup>b</sup> | S-MiXcan Cell-Type Specific Associations |  |  |  |
| --- | --- | --- | --- | --- | --- | --- | --- | --- | --- |
|  |  |  |  |  |  | Model | Overall P | Cell of Interest | Other Cells |
| 2p23.3 | NCOA1 | 2:24,490,992 | NDA, PD | — | — | Adipose | 2.1e-04 | 1.7e-04 (-) | 2.7e-04 (-) |
| 6q27 | THBS2-AS1 | 6:169,213,118 | NDA, PD | 1.5e-06 (+) | 8.7e-07 (+) | — | — | — | — |
| 8p11.23 | ZNF703 | 8:37,695,782 | NDA, PD | — | — | Epithelial | 3.9e-13 | 5.1e-08 (+) | 2.0e-13 (+) |
|  |  |  | NDA | — | — | Fibroblast | 2.8e-21 | 1.4e-21 (+) | 5.5e-10 (+) |
| 19q13.33 | SEC1P | 19:48,673,174 | DA | 8.1e-05 (+) | 2.7e-05 (+) | — | — | — | — |

a. Associations with overall breast cancer risk assessed using GWAS summary statistics for 133,384 case and 113,789 control women (Zhang 2020).
b. The most significant p-value among non-specific MiXcan models, which differ slightly due to randomness in the ensemble model estimation procedure.

### Independent replication

We evaluated associations of the 20 significant MD genes at 16 loci in an independent GWAS and TWAS of MD phenotypes in up to 27,900 European ancestry women [12] (**Supplementary Table 4 and 5**). Among the 11 genes at 10 loci that were examined in a prior TWAS using FUSION [15] tissue-level GReX models, 5 genes at 5 loci (*SV2A* at 1q21.2, *THBS2-AS1* at 6q27, *CCND1* at 11q13.3, *SEC1P* at 19q13.33, and *PELATON/SMIM25* at 20q13.13) had reported associations with the same MD phenotype (p<0.05) in the same direction as in the discovery sample. Replication analyses using independent GWAS summary statistics [12] and S-MiXcan to test associations using the same GReX prediction models as in the discovery TWAS showed that 5 additional genes at 4 loci (*POU3F3* at 2q12.1, *ZNF703* at 8p11.23, *LINC00445* at 13q13.3, and *NTN5* and *FUT2* at 19q13.33) were associated with the same MD phenotype (p<0.05) in the same direction. Altogether, associations of 10 MD genes at 8 loci were replicated in an independent population.

### Colocalization

Colocalization analyses [43] were performed based on GTEx v8 eQTL analyses of bulk mammary tissue from 329 men and women of European ancestry, and genome-wide associations in the study population of 24,158 European ancestry women (**Supplementary Table 6**). We found evidence (CLPP >0.01) that GWAS variants previously identified at 3 loci are causally associated with MD phenotypes through their effects on mammary tissue-level gene expression. Specifically, *FNBP1P1* expression mediates the association of DA and PD variants at 2p13.1, *ZNF703* expression mediates the association of NDA and PD variants at 8p11.23, and *PELATON/SMIM25* expression mediates the association of the DA variant at 20q13.13. Additionally, we found evidence of colocalization (CLPP >0.01) at a novel locus for NDA and PD at 6q27 whereby the rs3252 exonic variant in 2 (ENST00000796859 and ENST00000670651) out of 7 Ensembl transcripts of the *THBS2-AS1* antisense RNA is causally associated with expression levels and the NDA and PD phenotypes.

## DISCUSSION

Breast tissue composition is highly variable, not only between women, but also within the mammary tissue context where greater quantities of fibroglandular tissues increase breast cancer risk while healthy adipose tissues can play a protective role [5, 45]. We conducted the first cell-type-aware TWAS of mammographic density using MiXcan2, a new statistical tool for identifying genes associated with phenotypes through their predicted expression levels in specific cell types within the relevant tissue context. We identified 20 significant MD genes at 16 loci in 24,158 European ancestry women, including 6 novel genes at 6 independent loci not found by prior GWAS or TWAS of MD phenotypes. We discovered that one of the novel genes for NDA and PD, *THBS2-AS1* at 6q27, is also a novel breast cancer gene.

*THBS2-AS1* encodes the long non-coding RNA (lncRNA), thrombospondin-2 antisense RNA 1, which regulates the expression of thrombospondin-2. During mammary gland development, thrombospondin proteins play an important role in the differentiation of ductal epithelial cells and the formation of the mammary gland architecture [46]. Recent studies have shown that high tumor expression of THBS2 is a poor prognostic factor in breast cancer and other cancers, and that THBS2 may induce malignant transformation by enhancing epithelial-mesenchymal transition [47, 48]. *THBS2-AS1* expression in mammary tissue was significantly associated with decreased NDA and increased PD, as well as with increased breast cancer risk. The association of *THBS2-AS1* expression with nondense area was replicated in an independent study population [12]. Moreover, colocalization analyses revealed that the minor C-allele for exonic variant rs3252 is causally associated with increased mammary tissue *THBS2-AS1* expression, decreased nondense area and increased percent density, supporting the causal association of *THBS2-AS1* with breast phenotypes through its mammary tissue-level expression.

A second novel NDA gene, *CCND1* at 11q13.3, has been associated with breast cancer risk in a recent TWAS [38]. *CCND1* encodes cyclin D1, a regulatory subunit of the cyclin-dependent kinases CDK4 and CDK6 required for cell cycle G1/S transition. *CCND1* is an oncogene that is overexpressed in breast cancer [49]. Cyclin D1 is also required for normal mammary gland development during pregnancy [50]. We discovered that mammary tissue-level *CCND1* expression was significantly associated with decreased nondense area, and replicated this association in an independent population [12]. Cyclin D1 may decrease mammary adipogenesis by binding to the transcription factor PPARγ [49], which regulates adipocyte differentiation and has been associated with all three MD phenotypes and breast cancer risk in consistent directions by GWAS [6].

We also discovered 4 novel genes associated with DA (*VPS13D* at 1p36.22 and *AQP7* at 9p13.3), PD (*ZNF500* at 16p13.3), or both DA and PD (*POU3F3* at 2q12.1). *POU3F3* encodes a POU-domain containing protein that functions as a transcription factor, and may promote proliferation and inhibit apoptosis of cancer cells in triple-negative breast cancer [51]. *POU3F3* expression in mammary tissue was significantly associated with increased DA and PD, and both associations were independently replicated. *ZNF500* encodes the zinc finger protein 500 transcription factor and its expression in mammary tissue has been shown to decrease breast cancer cell proliferation [52]. Consistent with its known biology, we found that *ZNF500* expression in mammary tissue was associated with decreased PD. *AQP7* encodes aquaporin 7, a water and glycerol channel that regulates metabolic and signaling responses to cellular stress in breast cancer [53]. *AQP7* expression in mammary tissue was associated with decreased DA. *VPS13D* encodes the vacuolar protein sorting-associated protein 13D involved in protein trafficking. *VPS13D* expression in mammary fibroblasts was associated with increased DA.

In addition to discovering MD genes at 6 novel loci, our TWAS implicated candidate genes at 10 known GWAS loci that may be causally associated with MD phenotypes through their effects on gene expression levels in mammary tissue. Evidence from colocalization analyses showed that *FNBP1P1* associated with DA and PD loci at 2p13.1, *PELATON/SMIM25* associated with a DA locus at 20q13.13, and *ZNF703* associated with NDA and PD loci at 8p11.23 are likely to mediate the causal association of these GWAS loci with MD phenotypes. Additionally, TWAS analyses identified genes at 4 GWAS loci associated with MD phenotypes and breast cancer (*TBX15* at 1p12, *SV2A* at 1q21.2, *NCOA1* at 2p23.3, and *ADAMTS8* at 11q24.3) and 3 GWAS loci associated with MD phenotypes only (*ILK* and *ZNF215* at 11p15.4, *LINC00445* at 13q13.3, and *NTN5*, *SEC1P*, *FUT2* at 19q13.33).

MiXcan2 builds upon the MiXcan1 foundation for conducting cell-type-aware TWAS by expanding the set of genes and genetic predictors, and employing an ensemble modeling strategy to improve the robustness of GReX prediction models when the available bulk tissue training datasets are limited in size. MiXcan2 yields a higher proportion (∼60%) of nonspecific gene expression models than MiXcan1 (∼15%) enabling it to detect all 8 genes identified by standard tissue-level models developed using the same training set of female mammary tissue samples in this study. Additionally, MiXcan2 uniquely identified 12 genes not identified by standard approaches. This finding suggests that, with MiXcan2, it is no longer necessary to run both standard and cell-type-aware TWAS as recommended for MiXcan1, which tended to have lower power than PrediXcan in the absence of cell-type heterogeneity [17]. A recent method, scPrediXcan [54], develops cell-type-specific GReX prediction models from single-cell transcriptomic data by leveraging epigenomic regulatory databases. However, this approach is applicable only when an adequate number of individuals with single cell data is available. Currently, such data are typically obtainable for blood but are infeasible for female mammary tissues. As training datasets grow, future extensions of MiXcan2 may leverage both single-cell and bulk tissue data to enhance the power and interpretability of cell-type-aware TWAS for gene expression within the disease-relevant tissue context.

In summary, we conducted the first cell-type-aware TWAS of MD phenotypes in 24,158 European ancestry women using MiXcan2. We identified 20 MD genes at 16 loci tripling the total number from 8 genes at 7 loci [12] to 27 genes at 21 independent loci identified by TWAS in European ancestry women. We discovered that *THBS2-AS1* is a novel breast cancer gene that may increase breast cancer risk through its effects on decreasing NDA and increasing PD. Additionally, the *CCND1* oncogene may increase breast cancer risk in part through its effects on decreasing mammary adipogenesis and NDA. Few MD genes have been identified in women of non-European ancestry to date [13, 55, 56]. Future large multi-ancestry studies are needed to improve the power of gene discovery and to further elucidate the genetic etiology of mammographic density phenotypes and its relationship to breast cancer risk in all women.

## ACKNOWLEDGEMENTS

We are grateful to the Kaiser Permanente Northern California members who generously agreed to participate in the Research Program on Genes, Environment and Health. This study was supported by grants from the National Institutes of Health (R01CA23754, R01CA264987, R01CA244948, R21CA293419, U24CA210993, U24CA271114), Robert Wood Johnson Foundation, Ellison Medical Foundation, Wayne and Gladys Valley Foundation, and Kaiser Permanente National and Regional Community Benefit Programs. The authors acknowledge the computational resources and staff expertise provided by Scientific Computing at the Icahn School of Medicine at Mount Sinai. The BCAC breast cancer genome-wide association analyses were supported by the Government of Canada through Genome Canada and the Canadian Institutes of Health Research, the ‘Ministère de l’Économie, de la Science et de l’Innovation du Québec’ through Genome Québec and grant PSR-SIIRI-701, the National Institutes of Health (U19CA148065, X01HG007492), Cancer Research UK (C1287/A10118, C1287/A16563, C1287/A10710) and the European Union (HEALTH-F2-2009-223175 and H2020 633784 and 634935); all studies and funders are listed [30, 31]. The GTEx Project was supported by the Common Fund of theOffice of the Director of the National Institutes of Health, and by NCI, NHGRI, NHLBI, NIDA, NIMH, and NINDS. The results published here are in whole or part based upon data generated by the TCGA Research Network.

## FIGURES

**Supplementary Figure 1.**
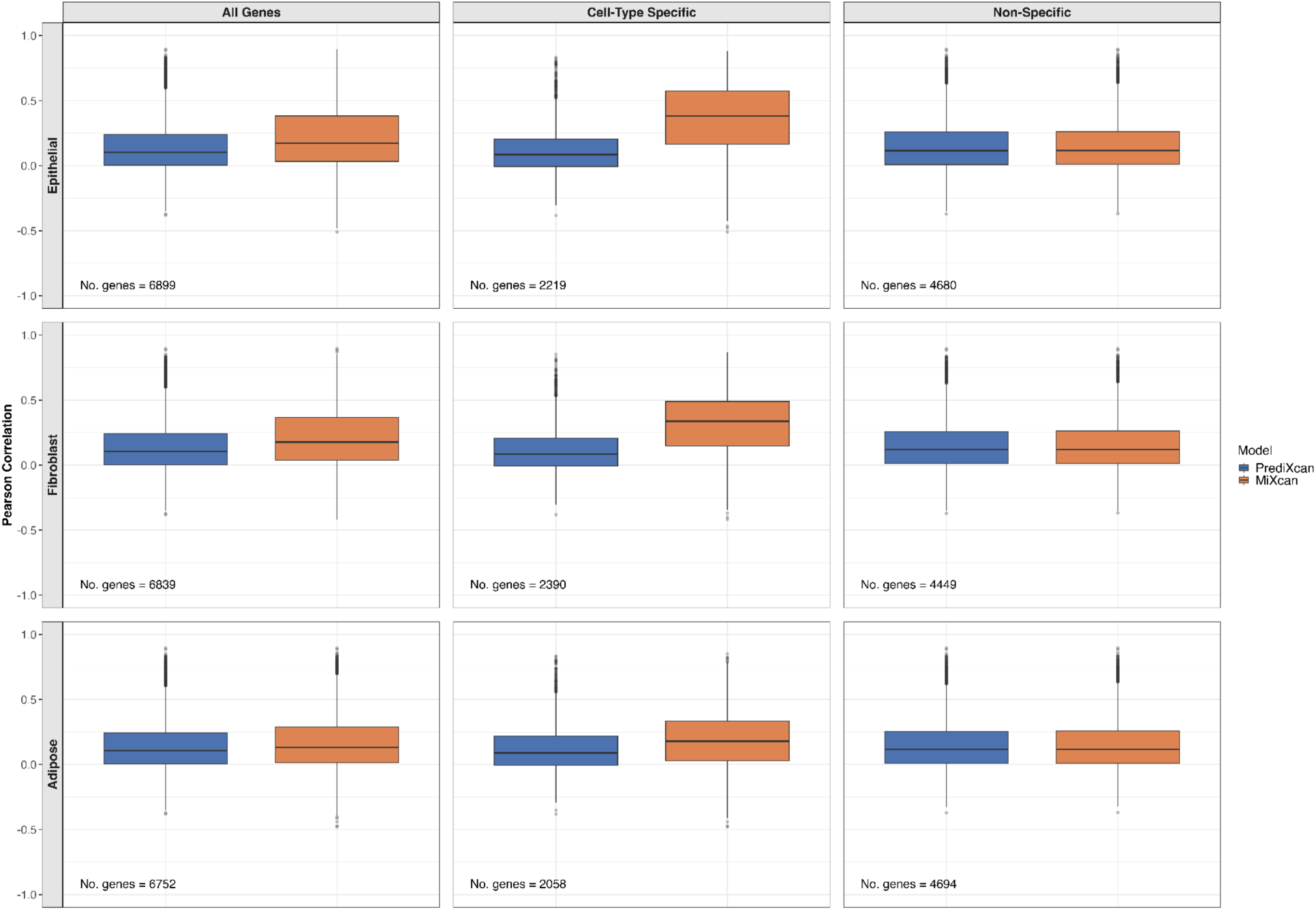
Pearson correlations between predicted and observed gene expression levels in 102 TCGA tumor-adjacent normal mammary tissue samples from European-ancestry women, for genes with both PrediXcan and MiXcan2 models.

**Supplementary Figure 2.**
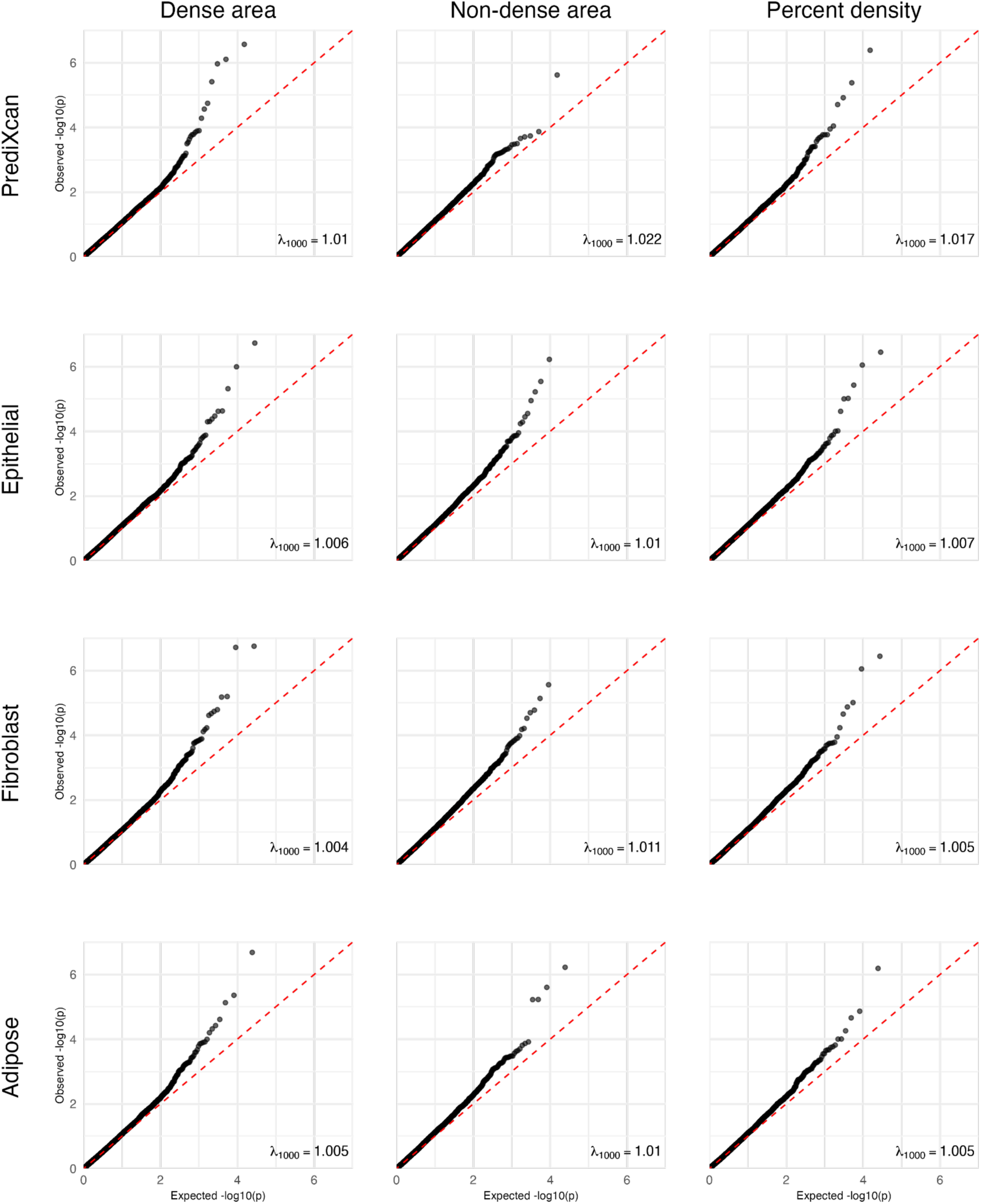
Quantile-quantile plots of observed vs. expected p-value distributions showed little evidence of inflated false-positive rates in transcriptome-wide analyses of mammographic dense area, nondense area, and percent density using PrediXcan, and MiXcan2 epithelial, fibroblast, and adipocyte models.

## REFERENCES

1. Sickles EA, d’Orsi CJ, Bassett LW, Appleton CM, Berg WA, Burnside ES: ACR BI-RADS Mammography. In: ACR BI-RADS Atlas, Breast Imaging Reporting and Data System. edn. Reston, Virginia: American College of Radiology; 2013.

2. Engmann NJ, Golmakani MK, Miglioretti DL, Sprague BL, Kerlikowske K, Breast Cancer Surveillance C: Population-Attributable Risk Proportion of Clinical Risk Factors for Breast Cancer. JAMA Oncol 2017, 3(9):1228–1236.

3. McCormack VA, dos Santos Silva I: Breast density and parenchymal patterns as markers of breast cancer risk: a meta-analysis. Cancer Epidemiol Biomarkers Prev 2006, 15(6):1159-1169.

4. Boyd NF, Guo H, Martin LJ, Sun L, Stone J, Fishell E, Jong RA, Hislop G, Chiarelli A, Minkin S et al: Mammographic density and the risk and detection of breast cancer. N Engl J Med 2007, 356(3):227–236.

5. Pettersson A, Graff RE, Ursin G, Santos Silva ID, McCormack V, Baglietto L, Vachon C, Bakker MF, Giles GG, Chia KS et al: Mammographic density phenotypes and risk of breast cancer: a meta-analysis. J Natl Cancer Inst 2014, 106(5).

6. Sieh W, Rothstein JH, Klein RJ, Alexeeff SE, Sakoda LC, Jorgenson E, McBride RB, Graff RE, McGuire V, Achacoso N et al: Identification of 31 loci for mammographic density phenotypes and their associations with breast cancer risk. Nat Commun 2020, 11(1):5116.

7. Arendt LM, Rudnick JA, Keller PJ, Kuperwasser C: Stroma in breast development and disease. Semin Cell Dev Biol 2010, 21(1):11–18.

8. Boyd NF, Dite GS, Stone J, Gunasekara A, English DR, McCredie MR, Giles GG, Tritchler D, Chiarelli A, Yaffe MJ et al: Heritability of mammographic density, a risk factor for breast cancer. N Engl J Med 2002, 347(12):886–894.

9. Stone J, Dite GS, Gunasekara A, English DR, McCredie MR, Giles GG, Cawson JN, Hegele RA, Chiarelli AM, Yaffe MJ et al: The heritability of mammographically dense and nondense breast tissue. Cancer Epidemiol Biomarkers Prev 2006, 15(4):612–617.

10. Ursin G, Lillie EO, Lee E, Cockburn M, Schork NJ, Cozen W, Parisky YR, Hamilton AS, Astrahan MA, Mack T: The relative importance of genetics and environment on mammographic density. Cancer Epidemiol Biomarkers Prev 2009, 18(1):102–112.

11. Lindstrom S, Thompson DJ, Paterson AD, Li J, Gierach GL, Scott C, Stone J, Douglas JA, dos-Santos-Silva I, Fernandez-Navarro P et al: Genome-wide association study identifies multiple loci associated with both mammographic density and breast cancer risk. Nat Commun 2014, 5:5303.

12. Chen H, Fan S, Stone J, Thompson DJ, Douglas J, Li S, Scott C, Bolla MK, Wang Q, Dennis J et al: Genome-wide and transcriptome-wide association studies of mammographic density phenotypes reveal novel loci. Breast Cancer Res 2022, 24(1):27.

13. Verma SS, Guare L, Ehsan S, Gastounioti A, Scales G, Ritchie MD, Kontos D, McCarthy AM, Penn Medicine B: Genome-Wide Association Study of Breast Density among Women of African Ancestry. Cancers (Basel) 2023, 15(10).

14. Gamazon ER, Wheeler HE, Shah KP, Mozaffari SV, Aquino-Michaels K, Carroll RJ, Eyler AE, Denny JC, Consortium GT, Nicolae DL et al: A gene-based association method for mapping traits using reference transcriptome data. Nat Genet 2015, 47(9):1091–1098.

15. Gusev A, Ko A, Shi H, Bhatia G, Chung W, Penninx BW, Jansen R, de Geus EJ, Boomsma DI, Wright FA et al: Integrative approaches for large-scale transcriptome-wide association studies. Nat Genet 2016, 48(3):245–252.

16. GTEx Consortium: The GTEx Consortium atlas of genetic regulatory effects across human tissues. Science 2020, 369(6509):1318-1330.

17. Song X, Ji J, Rothstein JH, Alexeeff SE, Sakoda LC, Sistig A, Achacoso N, Jorgenson E, Whittemore AS, Klein RJ et al: MiXcan: a framework for cell-type-aware transcriptome-wide association studies with an application to breast cancer. Nat Commun 2023, 14(1):377.

18. Banda Y, Kvale MN, Hoffmann TJ, Hesselson SE, Ranatunga D, Tang H, Sabatti C, Croen LA, Dispensa BP, Henderson M et al: Characterizing Race/Ethnicity and Genetic Ancestry for 100,000 Subjects in the Genetic Epidemiology Research on Adult Health and Aging (GERA) Cohort. Genetics 2015, 200(4):1285–1295.

19. Kvale MN, Hesselson S, Hoffmann TJ, Cao Y, Chan D, Connell S, Croen LA, Dispensa BP, Eshragh J, Finn A et al: Genotyping Informatics and Quality Control for 100,000 Subjects in the Genetic Epidemiology Research on Adult Health and Aging (GERA) Cohort. Genetics 2015, 200(4):1051–1060.

20. Emami NC, Cavazos TB, Rashkin SR, Cario CL, Graff RE, Tai CG, Mefford JA, Kachuri L, Wan E, Wong S et al: A Large-Scale Association Study Detects Novel Rare Variants, Risk Genes, Functional Elements, and Polygenic Architecture of Prostate Cancer Susceptibility. Cancer Res 2021, 81(7):1695–1703.

21. GTEx Consortium: Genetic effects on gene expression across human tissues. Nature 2017, 550(7675):204-213.

22. Byrska-Bishop M, Evani US, Zhao X, Basile AO, Abel HJ, Regier AA, Corvelo A, Clarke WE, Musunuri R, Nagulapalli K et al: High-coverage whole-genome sequencing of the expanded 1000 Genomes Project cohort including 602 trios. Cell 2022, 185(18):3426–3440 e3419.

23. Chang CC, Chow CC, Tellier LC, Vattikuti S, Purcell SM, Lee JJ: Second-generation PLINK: rising to the challenge of larger and richer datasets. Gigascience 2015, 4:7.

24. Petralia F, Krek A, Calinawan AP, Charytonowicz D, Sebra R, Feng S, Gosline S, Pugliese P, Paulovich AG, Kennedy JJ et al: BayesDeBulk: A Flexible Bayesian Algorithm for the Deconvolution of Bulk Tumor Data. bioRxiv 2023:2021.2006.2025.449763.

25. Aran D, Hu Z, Butte AJ: xCell: digitally portraying the tissue cellular heterogeneity landscape. Genome Biol 2017, 18(1):220.

26. Stegle O, Parts L, Durbin R, Winn J: A Bayesian framework to account for complex non-genetic factors in gene expression levels greatly increases power in eQTL studies. PLoS Comput Biol 2010, 6(5):e1000770.

27. Liu Y, Chen S, Li Z, Morrison AC, Boerwinkle E, Lin X: ACAT: A Fast and Powerful p Value Combination Method for Rare-Variant Analysis in Sequencing Studies. Am J Hum Genet 2019, 104(3):410–421.

28. Freedman ML, Reich D, Penney KL, McDonald GJ, Mignault AA, Patterson N, Gabriel SB, Topol EJ, Smoller JW, Pato CN et al: Assessing the impact of population stratification on genetic association studies. Nat Genet 2004, 36(4):388–393.

29. Devlin B, Roeder K: Genomic control for association studies. Biometrics 1999, 55(4):997–1004.

30. Michailidou K, Lindstrom S, Dennis J, Beesley J, Hui S, Kar S, Lemacon A, Soucy P, Glubb D, Rostamianfar A et al: Association analysis identifies 65 new breast cancer risk loci. Nature 2017, 551(7678):92-94.

31. Zhang H, Ahearn TU, Lecarpentier J, Barnes D, Beesley J, Qi G, Jiang X, O’Mara TA, Zhao N, Bolla MK et al: Genome-wide association study identifies 32 novel breast cancer susceptibility loci from overall and subtype-specific analyses. Nat Genet 2020, 52(6):572–581.

32. Feng H, Gusev A, Pasaniuc B, Wu L, Long J, Abu-Full Z, Aittomaki K, Andrulis IL, Anton-Culver H, Antoniou AC et al: Transcriptome-wide association study of breast cancer risk by estrogen-receptor status. Genet Epidemiol 2020, 44(5):442–468.

33. Jia G, Ping J, Shu X, Yang Y, Cai Q, Kweon SS, Choi JY, Kubo M, Park SK, Bolla MK et al: Genome-and transcriptome-wide association studies of 386,000 Asian and European-ancestry women provide new insights into breast cancer genetics. Am J Hum Genet 2022, 109(12):2185–2195.

34. Gao G, Fiorica PN, McClellan J, Barbeira AN, Li JL, Olopade OI, Im HK, Huo D: A joint transcriptome-wide association study across multiple tissues identifies candidate breast cancer susceptibility genes. Am J Hum Genet 2023, 110(6):950–962.

35. Li JL, McClellan JC, Zhang H, Gao G, Huo D: Multi-tissue transcriptome-wide association studies identified 235 genes for intrinsic subtypes of breast cancer. J Natl Cancer Inst 2024, 116(7):1105–1115.

36. Gao G, McClellan J, Barbeira AN, Fiorica PN, Li JL, Mu Z, Olopade OI, Huo D, Im HK: A multi-tissue, splicing-based joint transcriptome-wide association study identifies susceptibility genes for breast cancer. Am J Hum Genet 2024, 111(6):1100–1113.

37. Head ST, Dezem F, Todor A, Yang J, Plummer J, Gayther S, Kar S, Schildkraut J, Epstein MP: Cis-and trans-eQTL TWASs of breast and ovarian cancer identify more than 100 susceptibility genes in the BCAC and OCAC consortia. Am J Hum Genet 2024, 111(6):1084–1099.

38. Chang YH, Bresnahan ST, Taylor Head S, Harrison TA, Yu Y, Huff CD, Pasaniuc B, Lindstrom S, Bhattacharya A: Isoform-level analyses of 6 cancers uncover extensive genetic risk mechanisms undetected at the gene-level. Br J Cancer 2025, 133(6):874–885.

39. Ping J, Jia G, Cai Q, Guo X, Wang J, Tao R, Li B, Bauer JA, Xie Y, Ambs S et al: Multi-Ancestry Transcriptome-wide Association Studies Uncover New Insights into Breast Cancer Genetics and Biology. medRxiv 2025.

40. Zhu S, Song X: SMiXcan [Computer software]. https://githubcom/songxiaoyu/SMiXcan 2025.

41. Barbeira AN, Dickinson SP, Bonazzola R, Zheng J, Wheeler HE, Torres JM, Torstenson ES, Shah KP, Garcia T, Edwards TL et al: Exploring the phenotypic consequences of tissue specific gene expression variation inferred from GWAS summary statistics. Nat Commun 2018, 9(1):1825.

42. Cerezo M, Sollis E, Ji Y, Lewis E, Abid A, Bircan KO, Hall P, Hayhurst J, John S, Mosaku A et al: The NHGRI-EBI GWAS Catalog: standards for reusability, sustainability and diversity. Nucleic Acids Res 2025, 53(D1):D998–D1005.

43. Hormozdiari F, van de Bunt M, Segre AV, Li X, Joo JWJ, Bilow M, Sul JH, Sankararaman S, Pasaniuc B, Eskin E: Colocalization of GWAS and eQTL Signals Detects Target Genes. Am J Hum Genet 2016, 99(6):1245–1260.

44. Fernandez-Navarro P, Gonzalez-Neira A, Pita G, Diaz-Uriarte R, Tais Moreno L, Ederra M, Pedraz-Pingarron C, Sanchez-Contador C, Vazquez-Carrete JA, Moreo P et al: Genome wide association study identifies a novel putative mammographic density locus at 1q12-q21. Int J Cancer 2015, 136(10):2427–2436.

45. Pettersson A, Hankinson SE, Willett WC, Lagiou P, Trichopoulos D, Tamimi RM: Nondense mammographic area and risk of breast cancer. Breast Cancer Res 2011, 13(5):R100.

46. Pechoux C, Clezardin P, Dante R, Serre CM, Clerget M, Bertin N, Lawler J, Delmas PD, Vauzelle JL, Frappart L: Localization of thrombospondin, CD36 and CD51 during prenatal development of the human mammary gland. Differentiation 1994, 57(2):133-141.

47. Lin Y, Lin E, Li Y, Chen X, Chen M, Huang J, Guo W, Chen L, Wu L, Zhang X et al: Thrombospondin 2 is a Functional Predictive and Prognostic Biomarker for Triple-Negative Breast Cancer Patients With Neoadjuvant Chemotherapy. Pathol Oncol Res 2022, 28:1610559.

48. Ren Y, Ming R, Zuo A, Liu S, Ba Y, Zhang Y, Chen Y, Pan T, Luo P, Cheng Q et al: Cancer-associated fibroblasts drive lung adenocarcinoma progression via THBS2-mediated epithelial-mesenchymal transition. Oncogene 2025, 44(44):4284–4297.

49. Musgrove EA, Caldon CE, Barraclough J, Stone A, Sutherland RL: Cyclin D as a therapeutic target in cancer. Nat Rev Cancer 2011, 11(8):558–572.

50. Sicinski P, Weinberg RA: A specific role for cyclin D1 in mammary gland development. J Mammary Gland Biol Neoplasia 1997, 2(4):335–342.

51. Yang J, Meng X, Yu Y, Pan L, Zheng Q, Lin W: LncRNA POU3F3 promotes proliferation and inhibits apoptosis of cancer cells in triple-negative breast cancer by inactivating caspase 9. Biosci Biotechnol Biochem 2019, 83(6):1117–1123.

52. Ma X, Fan M, Yang K, Wang Y, Hu R, Guan M, Hou Y, Ying J, Deng N, Li Q et al: ZNF500 abolishes breast cancer proliferation and sensitizes chemotherapy by stabilizing P53 via competing with MDM2. Cancer Sci 2023, 114(11):4237–4251.

53. Dai C, Charlestin V, Wang M, Walker ZT, Miranda-Vergara MC, Facchine BA, Wu J, Kaliney WJ, Dovichi NJ, Li J et al: Aquaporin-7 Regulates the Response to Cellular Stress in Breast Cancer. Cancer Res 2020, 80(19):4071–4086.

54. Zhou Y, Adeluwa T, Zhu L, Salazar-Magana S, Sumner S, Kim H, Gona S, Nyasimi F, Kulkarni R, Powell JE et al: scPrediXcan integrates deep learning methods and single-cell data into a cell-type-specific transcriptome-wide association study framework. Cell Genom 2025, 5(5):100875.

55. Fejerman L, Ahmadiyeh N, Hu D, Huntsman S, Beckman KB, Caswell JL, Tsung K, John EM, Torres-Mejia G, Carvajal-Carmona L et al: Genome-wide association study of breast cancer in Latinas identifies novel protective variants on 6q25. Nat Commun 2014, 5:5260.

56. Mariapun S, Ho WK, Eriksson M, Tai MC, Mohd Taib NA, Yip CH, Rahmat K, Li J, Hartman M, Hall P et al: Evaluation of SNPs associated with mammographic density in European women with mammographic density in Asian women from South-East Asia. Breast Cancer Res Treat 2023, 201(2):237–245.

